# Chronic disease burden, not admission laboratory results, marks limb-threatening severity in acute lower-limb arterial thrombosis: a registry study of 288 patients from Kazakhstan

**DOI:** 10.64898/2026.07.11.26357804

**Authors:** Miras Mugazov, Yevgeniy Vruchinskiy, Alexey Fokin, Yermek Turgunov, Roza Yessimova, Kulash Nurseitova, Alina Ogizbayeva, Dinara Omertayeva

## Abstract

Acute lower-limb arterial thrombosis is a vascular emergency that often reaches internists and emergency physicians before a vascular specialist, and data from Central Asia are scarce. We asked which patient-level factors mark limb-threatening severity at first presentation, and whether routine admission laboratory tests add to that judgement. We studied a registry of 288 consecutive patients hospitalised with acute lower-limb arterial thrombosis between 2017 and 2022 at a tertiary centre in Karaganda, Kazakhstan. Severity was graded with the Savelyev classification, with grades IIB–IIIA defined as limb-threatening. Associations were tested by logistic regression with Firth-penalised and bootstrap sensitivity analyses, and predictors of tissue loss were modelled separately. Patients were mostly elderly men (median age 67 years, 72.2% male) with a heavy atherosclerotic burden (hypertension 74.0%, coronary disease 54.9%, diabetes 19.1%, prior lower-limb revascularisation 21.2% open and 14.6% endovascular). At presentation 142 (49.3%) had limb-threatening ischaemia and 31 (10.8%) tissue loss. Independent markers of limb-threatening severity were diabetes (adjusted odds ratio 2.15, 95% CI 1.02–4.55), hypertension (2.14, 1.05–4.37), tissue loss (2.55, 1.01–6.45) and lower body-mass index (0.71 per 5 kg/m², 0.52–0.97). Admission haematology, coagulation and metabolic values did not differ between severity groups (all p>0.05), and the chronic-disease model discriminated modestly (area under the curve 0.68). Low body-mass index (0.49 per 5 kg/m², p=0.005) and prior amputation (7.08, p=0.04) independently predicted tissue loss. In this cross-sectional analysis, limb-threatening severity was associated with the chronic vascular-metabolic phenotype rather than with the measured admission laboratory profile, which did not separate the severity groups; a leucocyte differential was not recorded, so the neutrophil-to-lymphocyte ratio could not be assessed. Low body weight was a consistent marker of more severe disease. At first contact the severity signal sat with the bedside assessment rather than the routine admission bloods examined here, a pattern that needs prospective testing.

## Introduction

Peripheral artery disease affects more than 110 million people, and most of them live in low- and middle-income countries, where the diagnosis tends to arrive late and the limb is often already at risk by presentation [1, 2]. Age-standardised prevalence has fallen in some high-income settings, yet the absolute number of affected people keeps rising as populations age and metabolic risk factors spread [3]. Central Asia sits inside this transition. Hypertension, diabetes and smoking remain poorly controlled across much of the region, and vascular care is concentrated in a small number of urban tertiary centres [4].

Acute lower-limb arterial thrombosis is the sharp end of this disease. Perfusion drops suddenly, the limb is threatened within hours, and the window for revascularisation is narrow [5]. Even with treatment the outlook is poor: in-hospital amputation reaches roughly 6% and one-year mortality has been reported as high as 41% [5]. The first clinician to see these patients is often an internist or an emergency physician rather than a vascular surgeon, and the bedside question is simple: which limb is in real trouble, and which patient can wait for an ordered work-up?

Severity at presentation drives that decision. The Rutherford categories, and in the post-Soviet region the Savelyev grades that map onto them, separate a viable limb from one that is immediately threatened [5, 6]. Less settled is which patient characteristics push a thrombosis towards the threatening end of that scale, and whether the blood tests routinely drawn on admission help. Inflammatory indices such as the neutrophil-to-lymphocyte ratio have drawn interest as prognostic markers after revascularisation [7, 8], but the European guideline panel concluded that no biomarker is established for routine prognostic use in this setting [5].

We had access to a six-year registry of patients admitted with acute lower-limb arterial thrombosis at a tertiary hospital in Karaganda, Kazakhstan. Our aim was twofold: to describe the clinical and laboratory profile of these patients in a region that rarely appears in the vascular literature, and to identify the factors that independently mark limb-threatening severity, with particular attention to whether admission laboratory tests carry any of that signal.

## Materials and methods

### Study design and setting

This was a single-centre, retrospective registry study reported in line with the STROBE recommendations for observational research [9]. The source population was every adult admitted with acute lower-limb arterial thrombosis to the vascular surgery service of a tertiary teaching hospital affiliated with Karaganda Medical University between January 2017 and December 2022, an unselected consecutive series with no exclusion by age, comorbidity or treatment. Records were held as an alphabetical patient listing; eight duplicate entries (identical name and date of birth) were identified and removed, leaving 288 unique patients for analysis. Data were extracted from the hospital’s medical records, which were accessed for research purposes between 01/04/2023 and 30/06/2023; being a consecutive series, the study had no formal sample-size calculation.

#### Ethics statement

The study used routinely collected, de-identified clinical data and was conducted in accordance with the Declaration of Helsinki. The protocol (“Prediction of the development of lower-limb arterial thrombosis in atherosclerosis”) was approved by the Local Bioethics Committee of Karaganda Medical University (registration No. 45 of 10 March 2023; committee decision of 27 March 2023, Extract from Protocol No. 7). The Local Bioethics Committee waived the requirement for individual informed consent for this retrospective analysis of anonymised records.

### Participants and variables

For each patient we recorded age, sex and body-mass index (BMI); the anatomical level of arterial occlusion (aorto-iliac, ilio-femoral, femoro-popliteal or popliteal-tibial); and the state of the contralateral limb, graded from patent to occluded. Cardiovascular comorbidity captured coronary artery disease, prior myocardial infarction, heart failure (NYHA class), arterial hypertension and its grade, prior stroke, carotid disease, diabetes mellitus and antiplatelet use. Previous vascular interventions (open revascularisation, lower-limb stenting, amputation) were noted, as was foot ulceration or necrosis at admission. Admission laboratory data included haemoglobin, erythrocyte, leucocyte and platelet counts, erythrocyte sedimentation rate, alanine and aspartate aminotransferases, total cholesterol, fasting glucose, activated partial thromboplastin time, prothrombin index, international normalised ratio and fibrinogen.

### Severity classification and outcomes

Ischaemia severity was graded by the treating surgeons at admission using the Savelyev (Pokrovsky) classification of acute limb ischaemia, standard practice across the post-Soviet region: grade IA (sensory loss / paraesthesia), IB (rest pain), IIA (motor paresis), IIB (motor paralysis) and IIIA (subfascial oedema). These grades map onto the Rutherford acute-ischaemia categories as follows: IA to category I (viable), IB and IIA to category IIa (marginally threatened), IIB to category IIb (immediately threatened) and IIIA to category III (irreversible/incipient) [5]. The primary outcome was limb-threatening ischaemia, defined a priori as grade IIB or IIIA: established motor paralysis or subfascial oedema marks the limb that is immediately threatened, for which emergent revascularisation rather than an ordered work-up is indicated. Patients with grade IA–IIA (intact or only partial motor function) formed the non-threatening comparison group. The secondary outcome was tissue loss, defined as foot ulceration or necrosis present at admission.

### Statistical analysis

Continuous variables are summarised as mean ± SD and median (interquartile range); categorical variables as counts and percentages. Implausible laboratory values, consistent with data-entry error, were set to missing using pre-specified physiological bounds before analysis. Severity groups were compared with the Mann–Whitney U test for continuous variables and the chi-square test (or Fisher’s exact test) for proportions; because the laboratory comparisons were exploratory, these p values are descriptive and were not adjusted for multiple testing. The multivariable logistic regression model was specified a priori and was not built by univariate screening. It included established vascular risk factors (age, sex, diabetes mellitus, arterial hypertension, coronary artery disease) together with markers of disease burden and anatomy (body-mass index, established tissue loss, severe contralateral disease, proximal occlusion level), entered together regardless of univariate significance. With 119 events and nine covariates the model held roughly 13 events per variable, above the conventional minimum. Age was scaled per 10 years and BMI per 5 kg/m² to give interpretable odds ratios; linearity of their relationship with the log-odds was checked and was acceptable. Discrimination was assessed by the area under the receiver-operating-characteristic curve. Because some covariate cells were small, the model was repeated with Firth’s penalised likelihood, and 95% confidence intervals were also derived from 2,000 bootstrap resamples. Predictors of tissue loss were modelled with Firth-penalised logistic regression given the lower event count. Analyses were complete-case; the pre-specified model variables were missing in under 6% of patients, so no imputation was performed. Because the model required all nine covariates, the complete-case multivariable analysis was fitted on 244 patients (119 limb-threatening events). A two-sided p<0.05 was considered significant. Computations used Python 3.12 (pandas, SciPy, statsmodels).

## Results

### Cohort characteristics

The 288 patients had a median age of 67 years (IQR 60–76); 208 (72.2%) were men. The cohort carried a dense burden of atherosclerotic and metabolic disease (Table 1; S1 Fig). Hypertension was present in 74.0%, coronary artery disease in 54.9% and chronic heart failure in 52.8%; 19.1% had diabetes and 32.6% were anaemic. Prior lower-limb revascularisation was common (open revascularisation 21.2%, stenting 14.6%; some patients had both), and 3.8% had a previous amputation. Despite this, only 63.9% were taking aspirin or another antiplatelet agent at admission, leaving roughly a third of an overtly high-risk population without basic secondary prevention. The occlusion was most often at the ilio-femoral (46.5%) or femoro-popliteal (41.7%) level.

**Table 1.** Baseline characteristics of 288 patients hospitalised with acute lower-limb arterial thrombosis.

| Characteristic | Value (n = 288) |
| --- | --- |
| **Demographics** |  |
| Age, years — median (IQR) | 67 (60–76) |
| Age, years — mean ± SD | 67.7 ± 11.8 |
| Male sex | 208 (72.2%) |
| Body-mass index, kg/m<sup>2</sup> — mean ± SD | 27.4 ± 4.7 |
| **Cardiovascular comorbidity** |  |
| Arterial hypertension | 213 (74.0%) |
| Grade-3 hypertension | 108 (37.5%) |
| Coronary artery disease | 158 (54.9%) |
| Chronic heart failure (any NYHA class) | 152 (52.8%) |
| Prior myocardial infarction (cardiosclerosis) | 57 (19.8%) |
| Diabetes mellitus | 55 (19.1%) |
| Prior stroke | 56 (19.4%) |
| Carotid artery disease | 25 (8.7%) |
| Anaemia | 94 (32.6%) |
| Antiplatelet therapy at admission | 184 (63.9%) |
| **Limb status and prior interventions** |  |
| Severe contralateral disease (occlusion / ≥80% stenosis) | 82 (28.6%) |
| Tissue loss (ulcer / necrosis) | 31 (10.8%) |
| Prior open revascularisation | 61 (21.2%) |
| Prior lower-limb stenting | 42 (14.6%) |
| Prior amputation | 11 (3.8%) |
| **Anatomical level of occlusion** |  |
| Aorto-iliac | 6 (2.1%) |
| Ilio-femoral | 134 (46.5%) |
| Femoro-popliteal | 120 (41.7%) |
| Popliteal-tibial | 28 (9.7%) |
| **Acute ischaemia grade (Savelyev)** |  |
| IA – sensory | 31 (10.8%) |
| IB – rest pain | 55 (19.1%) |
| IIA – paresis | 60 (20.8%) |
| IIB – paralysis | 134 (46.5%) |
| IIIA – subfascial oedema | 8 (2.8%) |
| Limb-threatening (IIB–IIIA) | 142 (49.3%) |

Values are n (%) unless otherwise stated. IQR, interquartile range; NYHA, New York Heart Association; SD, standard deviation.

### Severity at presentation

Almost half of the cohort, 142 patients (49.3%), reached hospital with a limb already immediately threatened (grade IIB in 134, IIIA in 8; Fig 1). A further 60 (20.8%) had motor paresis (IIA), and the remaining 86 presented with sensory loss or rest pain alone. Established tissue loss was present in 31 patients (10.8%).

**Fig 1.**
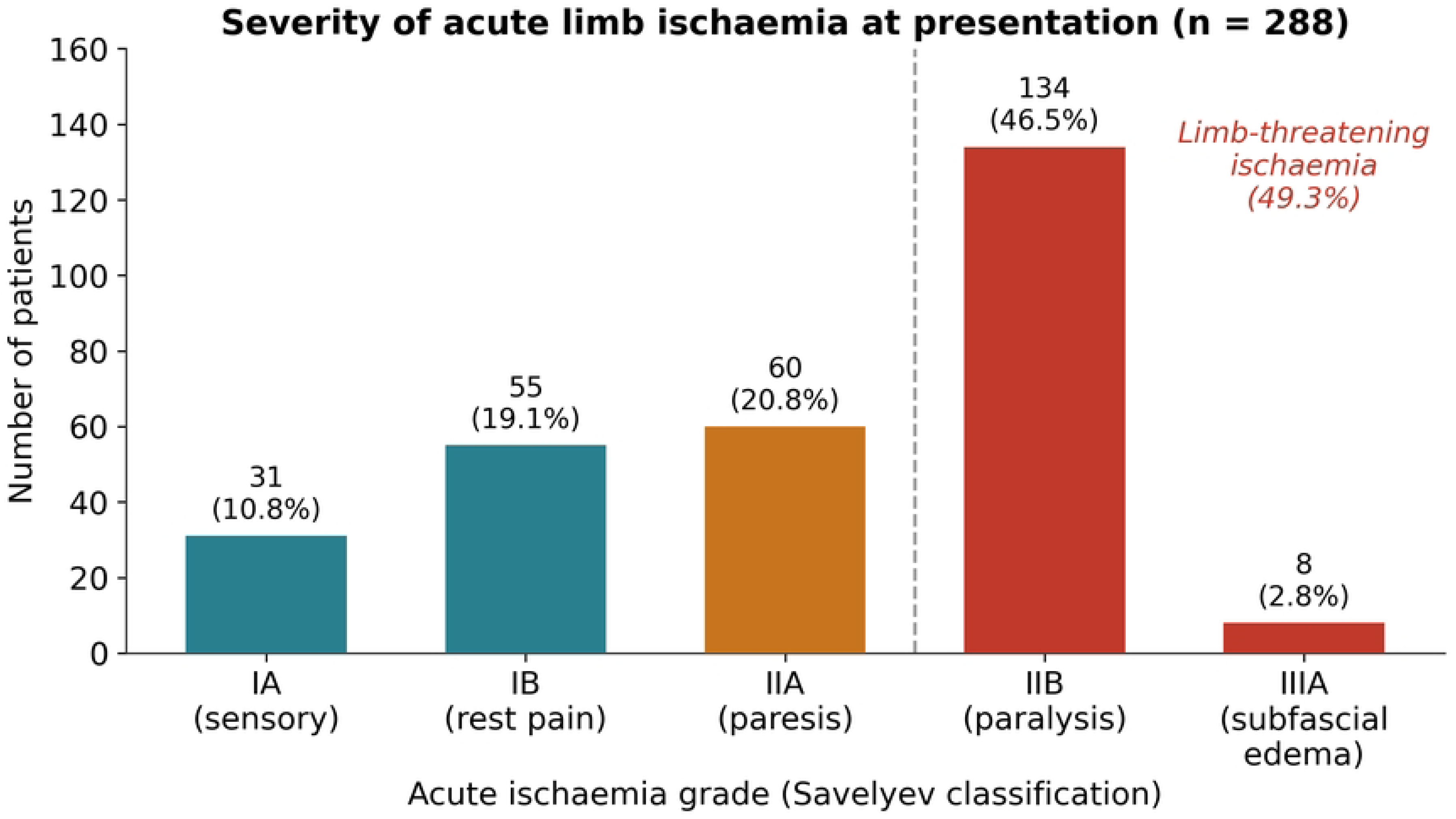
Distribution of acute ischaemia severity at presentation (Savelyev classification) among 288 patients. Grades IIB and IIIA, 49.3% of the cohort, denote an immediately threatened limb.

### Factors associated with limb-threatening ischaemia

On univariate comparison (Table 2), patients with limb-threatening ischaemia were older (mean 69.3 vs 66.1 years, p=0.025) and, counter-intuitively, leaner (BMI 26.6 vs 28.1 kg/m², p=0.011). They more often had established tissue loss (16.2% vs 5.5%, p=0.006) and severe contralateral disease (36.2% vs 21.2%, p=0.008), and the distribution of occlusion level differed between groups (p=0.038). Hypertension and erythrocyte sedimentation rate showed non-significant trends. None of the admission haematology, coagulation or metabolic values separated the two groups: haemoglobin, leucocyte and platelet counts, glucose, cholesterol, fibrinogen, prothrombin index, INR and aPTT were all statistically indistinguishable (Table 2).

**Table 2.** Comparison of patients with non-threatening versus limb-threatening ischaemia.

| Variable | Overall<br>(n=288) | Non-threatening<br>(n=146) | Limb-threatening<br>(n=142) | p |
| --- | --- | --- | --- | --- |
| ***Continuous, mean ± SD*** |  |  |  |  |
| Age, years | 67.7 ± 11.8 | 66.1 ± 12.5 | 69.3 ± 10.8 | 0.025* |
| Body-mass index, kg/m<sup>2</sup> | 27.4 ± 4.7 | 28.1 ± 4.7 | 26.6 ± 4.6 | 0.011* |
| Haemoglobin, g/L | 134 ± 20 | 134 ± 21 | 135 ± 20 | 0.86 |
| Leucocytes, ×10<sup>9</sup>/L | 9.6 ± 3.7 | 9.5 ± 3.3 | 9.7 ± 4.0 | 0.71 |
| Platelets, ×10<sup>9</sup>/L | 237 ± 88 | 229 ± 84 | 244 ± 91 | 0.11 |
| ESR, mm/h | 15.5 ± 13.5 | 14.6 ± 13.6 | 16.4 ± 13.3 | 0.079 |
| Glucose, mmol/L | 6.8 ± 3.1 | 6.7 ± 3.1 | 6.9 ± 3.1 | 0.58 |
| Total cholesterol, mmol/L | 4.6 ± 1.3 | 4.7 ± 1.4 | 4.6 ± 1.3 | 0.64 |
| Fibrinogen, g/L | 3.7 ± 1.6 | 3.6 ± 1.5 | 3.8 ± 1.7 | 0.46 |
| Prothrombin index, % | 89.1 ± 14.2 | 88.6 ± 14.4 | 89.6 ± 13.9 | 0.66 |
| INR | 1.2 ± 0.3 | 1.2 ± 0.3 | 1.2 ± 0.3 | 0.74 |
| aPTT, s | 36.2 ± 14.0 | 37.2 ± 15.7 | 35.2 ± 12.0 | 0.14 |
| ***Categorical, n (%)*** |  |  |  |  |
| Diabetes mellitus | 55 (19.1) | 25 (17.1) | 30 (21.1) | 0.48 |
| Arterial hypertension | 213 (74.0) | 101 (69.2) | 112 (78.9) | 0.082 |
| Coronary artery disease | 158 (54.9) | 80 (54.8) | 78 (54.9) | 1.00 |
| Chronic heart failure | 152 (52.8) | 72 (49.3) | 80 (56.3) | 0.28 |
| Tissue loss<br>(ulcer/necrosis) | 31 (10.8) | 8 (5.5) | 23 (16.2) | 0.006* |
| Severe contralateral<br>disease | 82 (28.6) | 31 (21.2) | 51 (36.2) | 0.008* |
| Anaemia | 94 (32.6) | 49 (33.6) | 45 (31.7) | 0.83 |
| Prior open revascularisation | 61 (21.2) | 30 (20.5) | 31 (21.8) | 0.90 |
| Prior amputation | 11 (3.8) | 5 (3.4) | 6 (4.2) | 0.97 |
| Occlusion level (overall $\chi^2$) | — | — | — | 0.038* |

*p* values from Mann–Whitney *U* (continuous) and chi-square (categorical) tests. \**p*<0.05. ESR, erythrocyte sedimentation rate; INR, international normalised ratio; aPTT, activated partial thromboplastin time.

### Multivariable analysis

In the multivariable model (Table 3, Fig 2; complete-case n=244, 119 events), five variables independently marked limb-threatening severity. Diabetes (adjusted OR 2.15, 95% CI 1.02–4.55), hypertension (2.14, 1.05–4.37) and established tissue loss (2.55, 1.01–6.45) each roughly doubled the odds, while every 5 kg/m² increase in BMI lowered them (0.71, 0.52–0.97). Diabetes and hypertension had not separated the groups on univariate testing (Table 2); they reached significance only after adjustment, which fits confounding by body habitus, since both cluster with the higher BMI that was itself protective. Severe contralateral disease sat at the edge of significance (1.82, 0.99–3.36), as did older age and female sex in the protective direction. The anatomical level of occlusion carried no independent weight once these factors were accounted for. The model discriminated modestly (AUC 0.68; Fig 3), with a likelihood-ratio p=0.0007. Firth-penalised estimates and bootstrap confidence intervals agreed closely with the primary model, so the findings were not an artefact of sparse cells (Table 3). For diabetes, hypertension and tissue loss the lower confidence limits sat close to 1.0, so these are best read as modest associations rather than strong effects.

**Fig 2.**
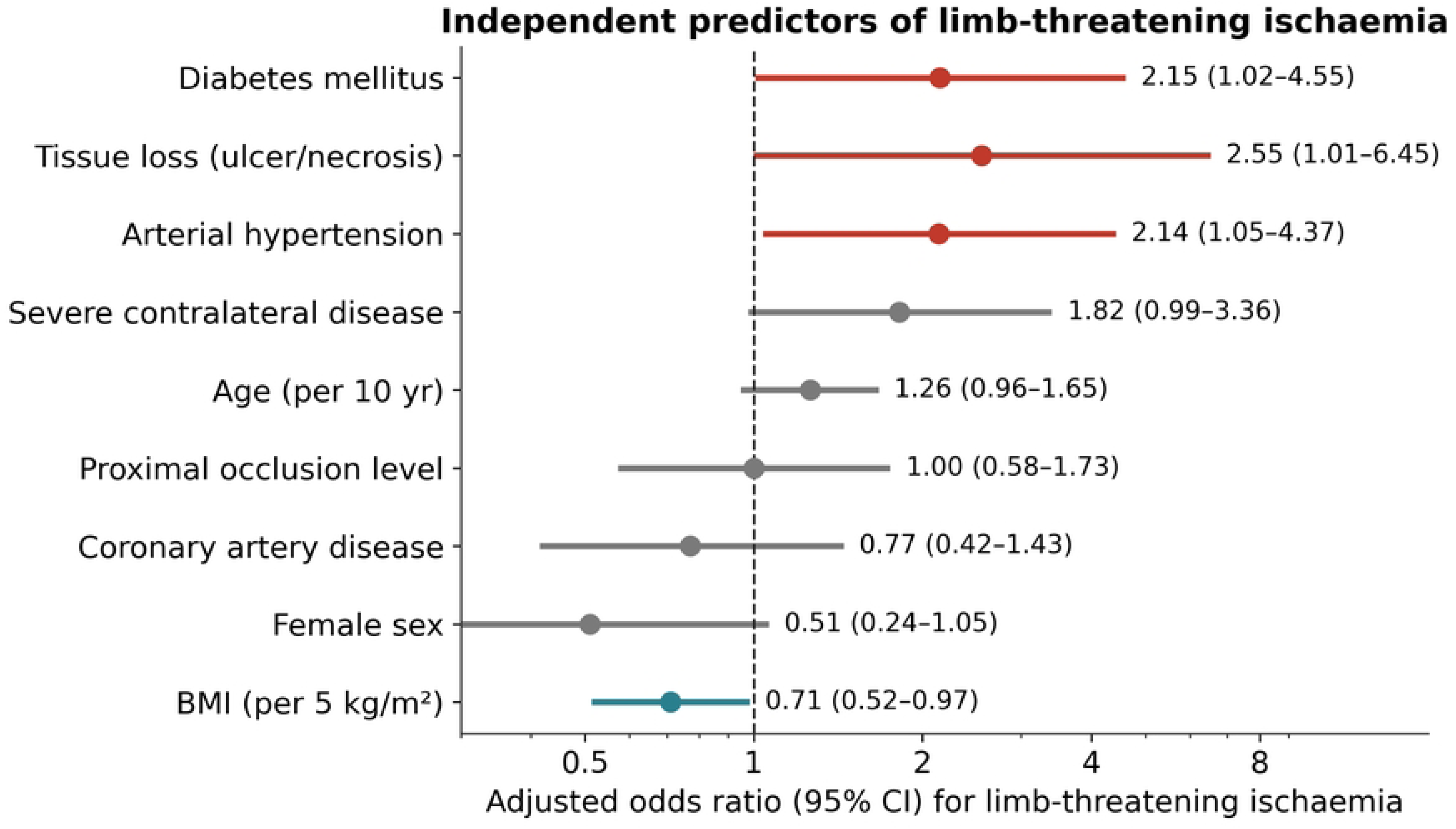
Adjusted odds ratios (95% CI) from the multivariable logistic regression for limb-threatening ischaemia. Red, significant risk factors; teal, significant protective factor; grey, non-significant. The vertical line marks an odds ratio of 1.

**Fig 3.**
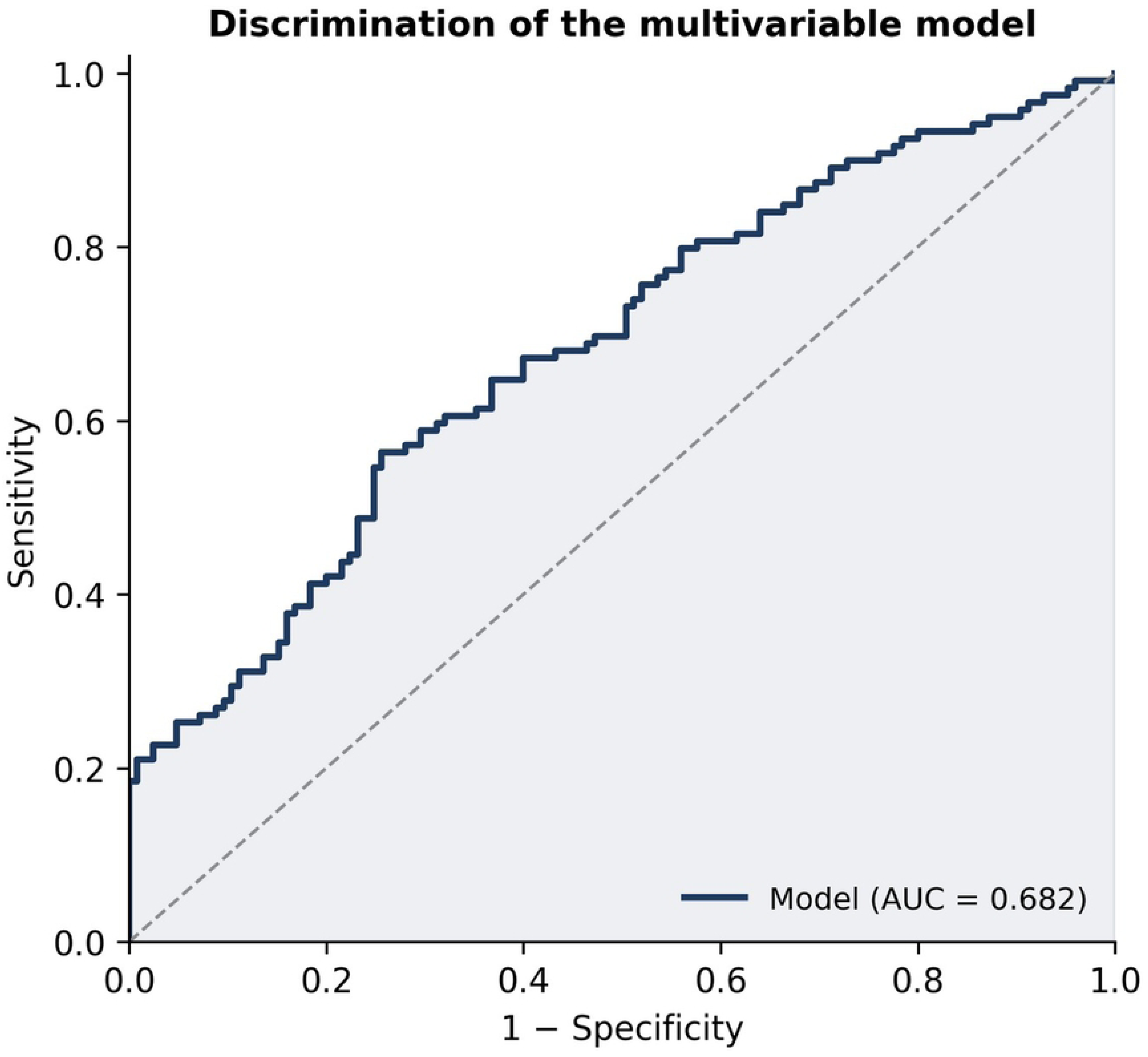
Receiver-operating-characteristic curve for the multivariable model (area under the curve 0.68).

**Table 3.** Multivariable logistic regression for limb-threatening ischaemia, with sensitivity analyses.

| Predictor | Adjusted OR (95% CI) | p | Firth OR / Bootstrap 95% CI |
| --- | --- | --- | --- |
| Diabetes mellitus | 2.15 (1.02–4.55) | 0.046 | 2.07 / 1.10–4.89 |
| Tissue loss (ulcer/necrosis) | 2.55 (1.01–6.45) | 0.047 | 2.41 / 1.10–8.46 |
| Arterial hypertension | 2.14 (1.05–4.37) | 0.037 | 2.07 / 1.09–4.84 |
| BMI (per 5 kg/m<sup>2</sup>) | 0.71 (0.52–0.97) | 0.032 | 0.72 / 0.49–0.98 |
| Severe contralateral disease | 1.82 (0.99–3.36) | 0.055 | 1.77 / 0.97–3.79 |
| Age (per 10 years) | 1.26 (0.96–1.65) | 0.095 | 1.24 / 0.95–1.72 |
| Female sex | 0.51 (0.24–1.05) | 0.068 | 0.52 / 0.22–1.03 |
| Coronary artery disease | 0.77 (0.42–1.43) | 0.41 | 0.78 / 0.39–1.49 |
| Proximal occlusion level | 1.00 (0.58–1.73) | 0.99 | 1.00 / 0.57–1.78 |

Complete-case analysis: $n=244$ , 119 events. Model AUC 0.68; likelihood-ratio $p=0.0007$ . The Firth column shows penalised-likelihood odds ratios; bootstrap interval from 2,000 resamples. OR, odds ratio; CI, confidence interval; BMI, body-mass index; AUC, area under the curve.

### Predictors of tissue loss

Tissue loss was modelled separately (S1 Table; Firth-penalised, n=243, 27 events). Two factors carried independent weight. Each 5 kg/m² of BMI again roughly halved the odds (OR 0.49, 95% CI 0.30–0.81, p=0.005), and a previous amputation carried a large independent risk (7.08, 1.06–47.4, p=0.04). Diabetes and limb-threatening ischaemia pointed the same way but did not reach significance in this smaller analysis. The low-BMI signal therefore ran through both outcomes.

## Discussion

Two results stand out in this registry. First, the severity of an acute lower-limb thrombosis at presentation was associated with the patient’s long-standing vascular and metabolic disease rather than with the numbers on the admission blood panel. Diabetes, hypertension, low body weight and existing tissue loss were the variables that independently flagged a limb at immediate risk. Second, the laboratory tests drawn on every one of these patients (the full blood count, the coagulation screen, glucose, cholesterol, fibrinogen) added little to the judgement of which limbs were threatened. For a clinician deciding how urgently to escalate, that is useful to know. The absent laboratory signal is worth dwelling on, because it runs against a common expectation. A high neutrophil-to-lymphocyte ratio predicts amputation and death after revascularisation for acute limb ischaemia in several series and in a recent meta-analysis [7, 8, 10], and inflammatory and coagulation markers are often assumed to mirror the acute event. Our data suggest they reflect outcome after intervention more than severity at the door. The European guideline reached a similar conclusion, finding no biomarker fit for routine prognostic use in this setting [5]. One caveat is real: our registry recorded total leucocyte counts but not the differential, so we could not compute the neutrophil-to-lymphocyte ratio itself. The crude counts did not discriminate, and because the leucocyte differential was unavailable and the sample size only moderate, our finding that the admission bloods add little should be read as applying to the routine values measured here rather than as evidence against any laboratory marker; we cannot exclude a role for the ratio.

The body-mass index finding is the most striking. Leaner patients had more severe ischaemia and more tissue loss, and the association survived adjustment in both models. At first glance this contradicts the well-known role of obesity as a driver of incident peripheral artery disease; in the ARIC cohort, for instance, a higher BMI predicted future hospitalisation for critical limb ischaemia [11]. The contradiction dissolves once the two questions are separated. Obesity helps cause the disease over decades, but once the disease is established it is the thin, often frail patient who fares worse, the so-called obesity, or lean, paradox seen across peripheral artery disease cohorts [12, 13, 14]. Low body weight in this population is rarely benign: it travels with sarcopenia, poor nutrition, chronic inflammation and depleted physiological reserve, and a meta-analysis of more than five million patients confirmed higher mortality in underweight patients with peripheral artery disease [15]. Our cross-sectional snapshot fits that pattern. A low BMI in someone presenting with acute limb thrombosis should read as a warning, not reassurance.

Diabetes and hypertension behaving as independent markers of severity is consistent with their established place in limb outcomes. Diabetes in particular roughly doubles the risk of amputation in peripheral artery disease and shifts disease towards the distal vessels that are hardest to revascularise [16]. The independent weight of existing tissue loss, and of prior amputation for the tissue-loss outcome, points the same way: these patients are well along their vascular journey, not at its start, which is exactly the population the global vascular guidelines on chronic limb-threatening ischaemia identify as needing urgent, structured, multidisciplinary care [6, 17]. Tissue loss here was a pre-existing foot wound recorded at admission, a marker of chronic limb-threatening ischaemia graded separately from the acute Savelyev category, so its association with a more severe acute presentation reflects accumulated disease rather than a restatement of the same measurement.

Two descriptive findings are worth flagging. Nearly half of the cohort reached hospital with an immediately threatened limb, which suggests late presentation is the rule here rather than the exception. And only about two-thirds were on any antiplatelet agent despite a comorbidity profile that all but mandates one. In a region where vascular services are thin and concentrated in cities [4], limbs arriving late and secondary prevention arriving incompletely probably matter more for population-level limb salvage than any refinement of in-hospital risk scoring.

### Clinical implications

For the internist or emergency physician, the practical message is that in this cross-sectional sample the severity signal at first contact sat with the history and the examination rather than with the admission bloods. A diabetic, hypertensive, low-weight patient with a foot wound or a hostile contralateral leg should be treated as high risk and referred urgently, however unremarkable the laboratory panel looks. A normal full blood count and coagulation screen should not be read as reassurance. The bloods are needed for peri-procedural safety, not for deciding who is threatened. These are associations from a single-centre, cross-sectional registry; they are meant to generate hypotheses, not to change practice before prospective validation.

### Strengths and limitations

The main strength is the cohort: 288 consecutive patients with acute lower-limb arterial thrombosis from a region almost absent from the vascular literature, with a broad clinical and laboratory dataset and severity graded by the treating surgeons at the bedside on admission. The agreement of the model across Firth and bootstrap analyses adds confidence. The limitations are those of a retrospective, single-centre registry. Causality cannot be inferred from a cross-sectional design, and unmeasured factors (symptom-to-door time, smoking status, thrombotic versus embolic mechanism, atrial fibrillation, renal function, the leucocyte differential) were not captured and may confound the associations. Severity was graded by the treating surgeons at the bedside, without blinding or a formal inter-rater check, which may introduce misclassification; and the embolic, in-situ thrombotic and graft-related mechanisms of occlusion, which carry different prognoses, were not distinguished. Tissue loss appeared both as a predictor in the primary model and as the secondary outcome; although the two were measured separately from the acute grade, their clinical overlap means the tissue-loss association in the primary model should be read as corroborative rather than independent. Age was derived from year of birth against the study midpoint, introducing minor misclassification. We had no follow-up, so the registry speaks to severity at presentation, not to amputation or survival. Missing data in some laboratory fields (platelets 21%, aPTT 16%) reduced the effective sample for those univariate comparisons; these high-missingness variables were not part of the pre-specified multivariable model, so a complete-case approach was reasonable and no imputation was performed. These gaps temper the conclusions but do not undermine the central, internally consistent pattern.

### Conclusions

In patients admitted with acute lower-limb arterial thrombosis in Kazakhstan, limb-threatening severity was associated with the chronic vascular-metabolic phenotype (diabetes, hypertension, low body-mass index and existing tissue loss) rather than with the admission laboratory profile, which did not separate the severity groups. Low body weight was a consistent marker of more severe disease across both severity and tissue loss, in line with the lean-paradox literature in established peripheral artery disease. Because the design is observational and cross-sectional and the model discriminated only modestly, these findings are hypothesis-generating: they are consistent with bedside assessment carrying more of the early severity signal than the routine admission bloods, and they point to late presentation and incomplete antiplatelet cover as the more likely places to look for regional gains in limb salvage. Whether any of this should change triage needs prospective testing.

## Acknowledgments

None.

## Author contributions

Data curation: Miras Mugazov, Roza Yessimova, Kulash Nurseitova.

Formal analysis: Roza Yessimova, Kulash Nurseitova.

Investigation: Miras Mugazov, Yevgeniy Vruchinskiy, Alina Ogizbayeva.

Methodology: Yevgeniy Vruchinskiy, Alexey Fokin, Yermek Turgunov.

Project administration: Dinara Omertayeva.

Supervision: Alexey Fokin, Yermek Turgunov, Dinara Omertayeva.

Validation: Alexey Fokin.

Visualization: Roza Yessimova, Kulash Nurseitova.

Writing – original draft: Miras Mugazov, Dinara Omertayeva.

Writing – review & editing: Miras Mugazov, Yevgeniy Vruchinskiy, Alexey Fokin, Yermek Turgunov, Roza Yessimova, Kulash Nurseitova, Alina Ogizbayeva, Dinara Omertayeva.

## Data availability

The data underlying the results presented in this study contain potentially identifying and sensitive patient information and are therefore subject to ethical restrictions imposed by the Bioethics Committee of Karaganda Medical University. Data cannot be shared publicly. De-identified data are available from the Bioethics Committee of Karaganda Medical University (Chair of the Local Bioethics Committee, Olga Aleksandrovna Visternichan) for researchers who meet the criteria for access to confidential data. Requests for data access may be sent to.

## Supporting information

S1 Table. Firth-penalised logistic regression for tissue loss (ulcer/necrosis).

S1 Fig. Prevalence of cardiovascular comorbidity, prior vascular interventions and limb findings across the whole cohort.

S2 Fig. Body-mass index (left) and age (right) by ischaemia severity. Boxes show median and interquartile range; points are individual patients.

